# ‘Self-weaning versus dose-adjusted caffeine regimens in preterm infants discharged home: a prospective cohort study’

**DOI:** 10.64898/2026.09.07.26361207

**Authors:** Shabina Ariff, Uzma Khan, Saima Fayyaz, Khubaib Abdullah, Ali Abbas, Siddiqa Ghani

## Abstract

**Objective:** To compare self-weaning and dose-adjusted caffeine regimens in preterm infants discharged home on caffeine for apnea of prematurity.

**Design:** Prospective cohort study.

**Setting:** Tertiary neonatal unit and outpatient clinics, Karachi, Pakistan, April 2023 to July 2025.

**Patients:** 204 preterm infants (<37 weeks’ gestation) discharged home on caffeine for apnea of prematurity; 94 self-weaning, 110 dose-adjusted.

**Interventions:** Self-weaning (absolute dose fixed, so mg/kg/day exposure declined with weight gain) versus dose-adjusted weaning (dose recalculated to current weight at each visit). Both were followed to 36 weeks postmenstrual age (PMA).

**Main outcome measures:** Proportion continuing caffeine beyond 36 weeks PMA and caregiver-reported apnea after discontinuation.

**Results:** Caffeine was continued beyond 36 weeks PMA in 68/94 self-weaning infants (72.3%) and 84/110 dose-adjusted infants (76.4%). No apnea was reported after discontinuation in either group, and all 204 infants survived to 36 weeks PMA. Apnea-related readmission on caffeine occurred in 1/94 (1.1%) versus 2/110 (1.8%). Weight at discontinuation was lower with self-weaning (1917 vs 2157 g); the dose-adjusted group was more premature (mean 30.0 vs 30.7 weeks). After adjustment for gestational age and birth weight, type of regimen was not associated with continuation beyond 36 weeks (adjusted OR 0.82, 95% CI 0.43 to 1.57) or time to discontinuation (adjusted HR 1.21, 95% CI 0.91 to 1.60).

**Conclusions:** Within a structured outpatient follow-up program, no post-discontinuation apnea was reported with either regimen, or apnea-related readmissions were infrequent. Self-weaning was not associated with adverse outcome and avoids repeated dose recalculation. A multicentre randomized trial with objective home monitoring is needed to determine the optimal dose and timing of discontinuation.

## INTRODUCTION

Apnea of prematurity (AOP) arises from immature central and peripheral chemoreceptor function and respiratory drive (1). It is defined as cessation of breathing for more than 20 seconds, or more than 10 seconds with bradycardia or desaturation, before 37 weeks’ gestation (2). The incidence of AOP increases with decreasing gestational age, affecting up to 85% of infants born before 34 weeks. In most cases, AOP resolves spontaneously by term-equivalent age (3,4)

The mainstay of pharmacologic therapy for AOP includes methylxanthines, with caffeine citrate preferred for its wider therapeutic index, longer half-life, and more predictable pharmacokinetics. (5,6) The Caffeine for Apnea of Prematurity (CAP) trial demonstrated that caffeine reduces apnea and is associated with shorter mechanical ventilation, less bronchopulmonary dysplasia (BPD), less severe retinopathy of prematurity, and better motor outcomes at five years. (7,8)

Despite widespread adoption of caffeine therapy in neonatal practice, optimal duration of treatment, timing of discontinuation, and criteria for hospital discharge remain subjects of debate. (9) Not many studies have evaluated structured approaches to continuing or weaning caffeine in the outpatient setting, even though clinical practice varies substantially across countries. In many high-income settings, infants remain hospitalized until caffeine has been stopped and apnea has resolved. (10)While this conservative approach prioritizes safety, it contributes to prolonged hospitalizations, increased healthcare costs, and additional psychosocial stress for families. In contrast, emerging evidence suggests that clinically stable infants may be safely discharged home on caffeine, provided adequate follow-up and monitoring are ensured, yet evidence guiding such practice remains limited. (11)

As healthcare systems increasingly seek strategies to reduce unnecessary NICU length of stay without compromising patient safety, outpatient caffeine management has gained attention. Two approaches are used: self-weaning, in which infants outgrow a fixed dose without active tapering, and dose-adjusted tapering, in which clinicians recalculate the dose at each visit. The theoretical ease and family-friendliness of self-weaning must be balanced against concerns about persistent subtherapeutic exposure or recurrence of apnea. At the same time, dose-adjusted tapering may allow more controlled discontinuation but requires more frequent clinical encounters. Given the lack of comparative evidence, it remains unclear which strategy optimally balances safety, feasibility, and resource use, particularly in low- and middle-income countries where home monitoring resources are limited, and follow-up adherence varies. This study aims to evaluate and compare the outcomes of preterm infants managed on self-weaning versus dose-adjusted caffeine regimens in an outpatient setting.

## METHODOLOGY

### Study Design and Setting

This prospective cohort study was conducted at the Neonatal Intensive Care Unit (NICU) and outpatient clinics of the Aga Khan University Hospital, Karachi, Pakistan, from April 2023 to July 2025. The study protocol was reviewed and approved by the institutional ethics review committee. Assignment of caffeine regimen was based solely on the clinical judgment of the attending neonatologist and did not involve any input from the research team. The patients were then tracked through a structured outpatient follow-up program via a dedicated weekly or biweekly neonatal clinic at our institution.

### Study Population

Preterm neonates born at less than 37 weeks’ gestation who received caffeine for apnea of prematurity (AOP) during their NICU stay and were discharged home on caffeine were eligible for inclusion. AOP was defined as above. Neonates with apnea attributable to sepsis, anemia of prematurity, or major congenital malformations, as well as those who died before discharge, discontinued caffeine before discharge, or had incomplete documentation, were excluded.

### Exposure and Caffeine Protocol

Eligible neonates were categorized into two groups based on the caffeine weaning strategy documented at follow-up visits. In the self-weaning group, the absolute caffeine dose remained fixed after discharge while the effective mg/kg/day exposure declined naturally with weight gain. In the dose-adjusted group, caffeine was recalculated according to current weight at each outpatient visit. Caffeine therapy was initiated in the NICU with a loading dose of 20 mg/kg of caffeine citrate, followed by a maintenance dose of 5–10 mg/kg/day (caffeine citrate) once daily. Dose adjustments during hospitalization were performed by the attending neonatologist based on daily weight measurements.

### Data Collection and Follow-Up

All study data were collected prospectively using a standardized proforma designed for the study. Neonatal data included gestational age, birth weight, Apgar scores, respiratory support requirements, comorbid diagnoses, caffeine dosing, and duration of therapy. Following discharge, all infants were followed through structured outpatient visits at the neonatal clinic until 36 weeks postmenstrual age (PMA). Primary analyses were anchored at 36 weeks PMA, but follow-up continued until caffeine discontinuation. At each visit, caregivers were asked about apneic episodes, feeding tolerance, weight gain, and any concerns. The corrected gestational age (CGA) at caffeine discontinuation and reasons for any readmissions were documented.

### Outcomes

The primary outcomes were the proportion of infants in whom caffeine was continued beyond 36 weeks PMA and the occurrence of caregiver-reported apnea after caffeine discontinuation, compared between the two groups. Secondary outcomes included CGA and weight at caffeine discontinuation, incidence of apnea at home while on caffeine, readmission after discharge for any cause and specifically due to apnea, caffeine-related adverse effects at home, and whether caffeine was restarted after discontinuation.

### Statistical Analysis

All analyses were performed in R (version 4.3.2; R Foundation for Statistical Computing). Categorical variables are summarised as n (%) and continuous variables as mean ± SD, except skewed durations, reported as median [IQR] with Wilcoxon rank-sum tests.Fisher’s exact test was used for all categorical comparisons to account for low expected cell counts in rare-event categories, and the independent-samples t-test was used for continuous variables. To address baseline imbalances, multivariable logistic regression was performed to estimate the adjusted odds ratio (aOR) for continuing caffeine beyond 36 weeks, with gestational age at birth and birth weight included as continuous covariates. Time-to-event analysis was conducted using the Kaplan-Meier method with log-rank testing via the survival and survminer packages, and a multivariable Cox proportional-hazards model was fitted to estimate the adjusted hazard ratio (aHR) for caffeine discontinuation. The proportional hazards assumption was evaluated using the Schoenfeld residuals test. Tables were exported to Word using the flextable and officer packages. All reported p-values were two-tailed, with significance set at p < 0.05.

## RESULTS

### Study Population and Baseline Characteristics

A total of 10,747 babies were delivered at Aga Khan University from April 2023 to July 2025. Of 2,666 preterm deliveries, 1,203 were admitted to the NICU, and 482 met screening criteria. After 278 exclusions (Figure 1), 204 infants were analyzed: 94 self-weaning and 110 dose-adjusted.The baseline maternal and infant characteristics are detailed in Table 1. Infants in the dose-adjusted group were more premature (30.0 ± 1.8 vs 30.7 ± 1.8 weeks, p = 0.007) and more often received surfactant; birth weight, sex, and Apgar scores were comparable (Table 1).

**Figure 1:**
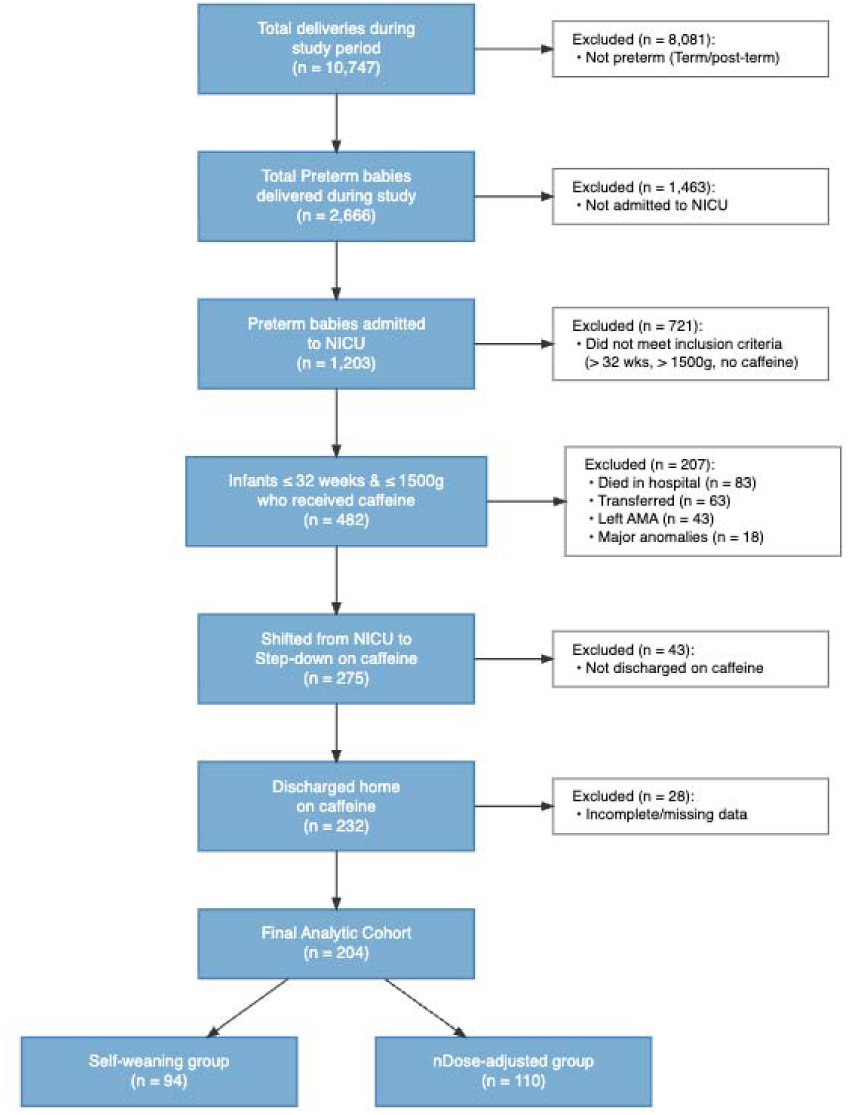
Number of Infants Eligible for Study and Categorized by Outpatient Caffeine Management Strategy.

**Table 1.** Maternal Characteristics and Infant Demographics (N = 204)

|  | Characteristic<sup>1</sup> | Self-Weaning (N = 94)<sup>1</sup> | Dose-Adjusted (N = 110)<sup>2</sup> | *p*-value |
| --- | --- | --- | --- | --- |
| Maternal Characteristics | Antenatal Steroids | 84 (89.4%) | 83 (75.5%) | 0.011 |
|  | Mode of Delivery |  |  | >0.999 |
|  | C-Section | 89 (94.7%) | 103 (93.6%) |  |
| Infant Demographics | Gestational Age at Birth (weeks) | 30.7 ± 1.8 | 30.0 ± 1.8 | 0.007 |

| Characteristic<sup>1</sup> | Self-Weaning (N = 94)<sup>1</sup> | Dose-Adjusted (N = 110)<sup>2</sup> | *p*-value |
| --- | --- | --- | --- |
| Birth Weight (grams) | 1,306.4 ± 301.7 | 1,259.6 ± 231.2 | 0.221 |
| Gender |  |  | >0.999 |
| Male | 49 (52.1%) | 58 (52.7%) |  |
| Female | 45 (47.9%) | 52 (47.3%) |  |
| APGAR at 1 Minute | 6.9 ± 1.6 | 7.1 ± 1.5 | 0.309 |
| APGAR at 5 Minutes | 8.6 ± 1.1 | 8.6 ± 0.8 | 0.774 |

<sup>1</sup>n (%); Mean ± SD
<sup>2</sup>Fisher's exact test; Welch Two Sample t-test
Values are presented as Mean ± SD for continuous variables and n (%) for categorical variables. P-values from independent t-test (continuous) and Fisher's exact test (categorical).

### Hospital Course

The hospital course and complication profile are presented in Table 2. Surfactant was administered more frequently in the dose-adjusted group (65.1% vs 46.8%, p = 0.011), and the length of hospital stay was significantly longer (28.5 ± 17.0 vs. 23.8 ± 12.5 days, p = 0.024). The median duration of mechanical ventilation was also longer in the dose-adjusted group (2.0 vs. 1.0 days, p = 0.028). CGA and weight at discharge were similar between groups (Table 2). Apnea during hospitalization, RDS, and NEC did not differ.

**Table 2.** Hospital Course and Complications (N = 204)

| Characteristic<sup>1</sup> | Self-Weaning (N = 94)<sup>1</sup> | Dose-Adjusted (N = 110)<sup>2</sup> | *p*-value |
| --- | --- | --- | --- |
| **Hospital Course** |  |  |  |
| Surfactant Administered | 44 (46.8%) | 71 (65.1%) | 0.011 |
| Apnea During Hospital Stay | 59 (62.8%) | 77 (70.0%) | 0.299 |
| Non-Invasive Ventilation | 91 (96.8%) | 108 (98.2%) | 0.663 |
| Duration of NIV (Days) |  |  | 0.037 |

|  | Characteristic<sup>1</sup> | Self-Weaning | Dose-Adjusted (N | *p*- |
| --- | --- | --- | --- | --- |
|  |  | (N = 94)<sup>1</sup> | = 110)<sup>2</sup> | value |
|  | Median [Q1 – Q3] | 10.0 [4.0 – 18.0] | 12.0 [ 7.0 – 22.0] |  |
|  | Mechanical Ventilation | 48 (51.1%) | 70 (63.6%) | 0.088 |
|  | Duration of Mechanical Vent (Days) |  |  | 0.028 |
|  | Median [Q1 – Q3] | 1.0 [0.0 – 3.0] | 2.0 [0.0 – 4.0] |  |
|  | Length of Hospital Stay (days) | 23.8 ± 12.5 | 28.5 ± 17.0 | 0.024 |
|  | CGA at Discharge (weeks) | 34.2 ± 1.3 | 34.1 ± 1.5 | 0.637 |
|  | Weight at Discharge (grams) | 1,509.5 ± 236.9 | 1,515.5 ± 252.8 | 0.861 |
|  | Type of Feed (24h Pre-Discharge) |  |  | 0.075 |
|  | Exclusive Breast Milk | 63 (67.0%) | 64 (58.2%) |  |
|  | Mixed Feeding | 30 (31.9%) | 38 (34.5%) |  |
|  | Formula Only | 1 (1.1%) | 8 (7.3%) |  |
| Complications During Hospital Stay | RDS | 74 (78.7%) | 96 (87.3%) | 0.132 |
|  | BPD | 0 (0.0%) | 5 (4.5%) | 0.063 |
|  | NEC | 4 (4.3%) | 6 (5.5%) | 0.756 |

| Characteristic<sup>1</sup> | Self-Weaning<br>(N = 94)<sup>1</sup> | Dose-Adjusted (N<br>= 110)<sup>2</sup> | *p*-<br>value |
| --- | --- | --- | --- |
| IVH | 0 (0.0%) | 5 (4.5%) | 0.063 |

<sup>1</sup>n (%); Mean ± SD
<sup>2</sup>Fisher's exact test; Welch Two Sample t-test

Values are presented as Mean ± SD for continuous variables and n (%) for categorical variables. P-values from independent t-test (continuous) and Fisher’s exact test (categorical).

Values are presented as Mean ± SD for continuous variables and n (%) for categorical variables. P-values from independent t-test (continuous) and Fisher’s exact test (categorical). RDS = Respiratory Distress Syndrome; BPD = Bronchopulmonary Dysplasia; NEC = Necrotizing Enterocolitis; IVH = Intraventricular Hemorrhage.

### Primary Outcomes

Caffeine was continued beyond 36 weeks PMA in 68/94 (72.3%) self-weaning and 84/110 (76.4%) dose-adjusted infants (p = 0.524). No apnea was reported after discontinuation in either group, and all 204 infants survived to 36 weeks PMA (Table 3).

**Table 3.** Primary and Secondary Clinical Outcomes (N = 204)

| Outcome<sup>1</sup> | Self-Weaning<br>(N = 94)<sup>1</sup> | Dose-Adjusted<br>(N = 110)<sup>2</sup> | *p*-<br>value |
| --- | --- | --- | --- |
| **Primary Outcomes at**<br>**36 Weeks PMA** |  |  |  |
| Caffeine Continued Beyond<br>36 Weeks | 68 (72.3%) | 84 (76.4%) | 0.524 |
| Caffeine Discontinued Before<br>or At 36 Weeks | 26 (27.7%) | 26 (23.6%) |  |

|  | Outcome<sup>1</sup> | Self-Weaning<br>(N = 94)<sup>1</sup> | Dose-Adjusted<br>(N = 110)<sup>2</sup> | *p*-<br>value |
| --- | --- | --- | --- | --- |
| Secondary Outcomes | CGA at Caffeine Discontinuation |  |  | 0.424 |
|  | ≤ 34.0 weeks | 3 (3.2%) | 2 (1.8%) |  |
|  | 34.1–36.0 weeks | 23 (24.5%) | 24 (21.8%) |  |
|  | 36.1–38.0 weeks | 55 (58.5%) | 60 (54.5%) |  |
|  | 38.1–40.0 weeks | 12 (12.8%) | 18 (16.4%) |  |
|  | > 40 weeks | 1 (1.1%) | 6 (5.5%) |  |
|  | Weight at Caffeine Discontinuation (grams) | 1,917.1 ± 471.6 | 2,156.8 ± 468.9 | 0.002 |
|  | Caffeine Restarted After Discontinuation | 0 (0.0%) | 2 (1.8%) | 0.501 |
|  | Apnea Reported at Home (On Caffeine) | 5 (5.3%) | 9 (8.2%) | 0.580 |
|  | Readmission After Discharge (Any Cause) | 8 (8.5%) | 16 (14.5%) | 0.199 |
|  | Readmission Due to Apnea (On Caffeine) | 1 (1.1%) | 2 (1.8%) | >0.999 |

<sup>1</sup>n (%); Mean ± SD
<sup>2</sup>Fisher's exact test; Welch Two Sample t-test

Values are presented as Mean ± SD for continuous variables and n (%) for categorical variables. P-values from independent t-test (continuous) and Fisher’s exact test (categorical).

CGA = Corrected Gestational Age; PMA = Postmenstrual Age. Weight at caffeine discontinuation was derived from the outpatient visit weight closest to the documented discontinuation date. No apneic episodes were reported after caffeine discontinuation in either group, and no readmissions due to apnea occurred after caffeine was discontinued.

### Secondary Outcomes

The distribution of CGA at caffeine discontinuation was statistically similar across groups (p = 0.424), with the majority of infants in both groups discontinuing caffeine between **36.1-38.0 weeks** (Table 3). Weight at caffeine discontinuation was significantly lower in the self-weaning group (1,917.1 ± 471.6 vs. 2,156.8 ± 468.9 g, p = 0.002). Apnea at home while on caffeine was infrequent in both groups (5.3% vs 8.2%, p = 0.580). Readmission due to apnea while on caffeine occurred in 1 infant (1.1%) in the self-weaning group and 2 infants (1.8%) in the dose-adjusted group (p > 0.999), and no readmissions due to apnea occurred after caffeine was discontinued. Caffeine was not restarted in any infant in the self-weaning group, and in 2 infants (1.8%) in the dose-adjusted group (p = 0.501). Very few non-specific caffeine-related adverse effects were documented at home. Parental discontinuation of caffeine before the scheduled visit was observed in 11.7% and 9.1% of infants in the self-weaning and dose-adjusted groups, respectively (p = 0.646).

### Adjusted Analyses

Given the significant baseline difference in gestational age and the longer hospital stay in the dose-adjusted group, multivariable models were constructed to determine whether the caffeine regimen independently influenced outcomes after adjustment for prematurity severity. In the logistic regression model, the self-weaning regimen was not independently associated with higher odds of continuing caffeine beyond 36 weeks compared with the dose-adjusted regimen (aOR 0.82, 95% CI 0.43–1.57, p = 0.551), and neither gestational age (aOR 1.04, 95% CI 0.82–1.31, p = 0.767) nor birth weight (aOR 1.00, 95% CI 1.00–1.00, p = 0.328) were significant predictors (Table 4). Kaplan-Meier analysis of outpatient caffeine duration revealed that the self-weaning group demonstrated a trend toward earlier discontinuation, with a median outpatient duration of approximately 2.7 weeks compared with 3.3 weeks in the dose-adjusted group (Figure 2). In the Cox proportional-hazards model, which satisfied the proportional hazards assumption (Schoenfeld global test p = 0.967), infants in the self-weaning group had a numerically higher adjusted hazard of caffeine discontinuation (aHR 1.21, 95% CI 0.91–1.60, p = 0.188), suggesting a trend toward faster weaning that was not statistically significant after controlling for gestational age and birth weight. However, higher gestational age at birth (aHR 1.27, 95% CI 1.14–1.41, p < 0.001) and birth weight (aHR 1.00, 95% CI 1.00–1.00, p = 0.024) were significantly associated with a faster rate of caffeine discontinuation (Table 5).

**Figure 2:**
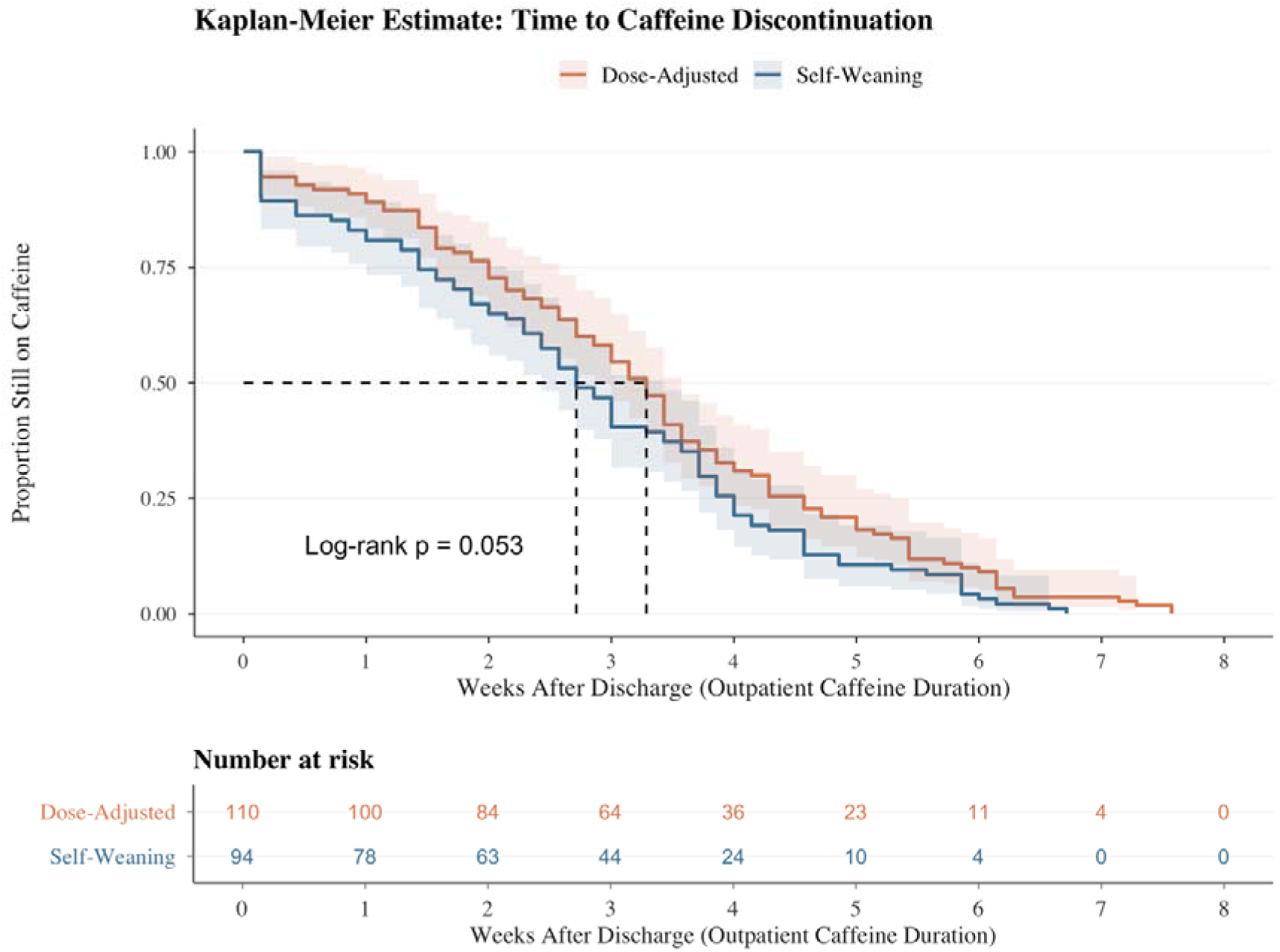
Kaplan-Meier survival curve comparing time to caffeine discontinuation between Dose-Adjusted and Self-Weaning groups over 8 weeks after discharge

**Table 4.** Multivariable Logistic Regression – Adjusted Odds of Continuing Caffeine Beyond 36 Weeks PMA (N = 204)

| Variable | aOR | 95% CI | p-value |
| --- | --- | --- | --- |
| **Caffeine Regimen** |  |  |  |
| Dose-Adjusted | — | — |  |
| Self-Weaning | 0.82 | 0.43, 1.57 | 0.551 |
| **Gestational Age (weeks)** | 1.04 | 0.82, 1.31 | 0.767 |
| Birth Weight (grams) | 1.00 | 1.00, 1.00 | 0.328 |

**Table 5.** Cox Proportional-Hazards Regression – Adjusted Hazard of Caffeine Discontinuation.

**Discontinuation\*\***
| Characteristic | aHR | 95% CI | p-value |
| --- | --- | --- | --- |
| **Caffeine Regimen** |  |  |  |
| Dose-Adjusted | — | — |  |
| Self-Weaning | 1.21 | 0.91, 1.60 | 0.188 |
| **Gestational Age (weeks)** | 1.27 | 1.14, 1.41 | <0.001 |
| **Birth Weight (grams)** | 1.00 | 1.00, 1.00 | 0.024 |

Abbreviations: CI = Confidence Interval, HR = Hazard Ratio

## DISCUSSION

In this prospective cohort of 204 preterm infants discharged home on caffeine, no breakthrough apneic episodes were reported in either the self-weaning or the dose-adjusted group after caffeine discontinuation. Apnea-related readmissions during caffeine therapy were rare in both groups (1.1% vs 1.8%, p>0.999). Although infants in the dose-adjusted group were significantly more premature at baseline (gestational age 30.0 ± 1.8 vs 30.7 ± 1.8 weeks, p=0.007) and had longer hospital stays, outpatient clinical outcomes did not differ between regimens after adjustment for these baseline differences. To our knowledge, this is one of the first prospective comparative datasets from a low- and middle-income country (LMIC) tertiary neonatal unit, most previous reports having described only a single regimen in high-income populations. (11,12)

In high-income settings, infants typically remain inpatient until 37–40 weeks PMA, off caffeine and apnea-free for 5–8 days. (10). This is protective but prolongs hospitalization, extends maternal-infant separation with attendant nosocomial risk, and increases cost. The present cohort, by comparison, was discharged at a mean corrected gestational age of approximately 34 weeks (substantially earlier than HIC norms), a pattern made feasible by outpatient caffeine continuation under structured neonatal clinic follow-up. Several factors drive this earlier discharge in LMIC settings, mainly limited NICU bed capacity relative to demand, limited kangaroo mother care (KMC) , the direct and indirect financial burden on families, and other social factors.(13) The home monitoring infrastructure that HIC protocols implicitly assume (apnea monitors, continuous pulse oximetry, community midway follow-ups, and rapid 24-hour access to emergency transport) was largely unavailable to these families, principally because the trial was unfunded.

Outpatient caffeine continuation is increasingly accepted as a practical solution, yet formal protocols remain inconsistent owing to the limited evidence base. The WHO currently offers no explicit guidance on home caffeine continuation in resource-limited settings, leaving individual units to formulate their own discharge frameworks and protocols. Self-weaning strategy may offer greater practical advantages in LMIC settings. As infants gain weight, the effective caffeine dose decreases without requiring dose adjustments, reducing the incremental cost of therapy while avoiding repeated weight-based dose recalculations. This lessens dependence on reliable weighing equipment, trained healthcare personnel, and accurate dose calculations, all of which may be limited in resource-constrained settings.

The optimal timing of caffeine discontinuation remains poorly characterized, and norms vary considerably across centers. The recent Cochrane review by Urru et al. reached the same conclusion, noting that included studies clustered across a broad PMA range and rating the certainty of evidence as low to moderate. (10) Most reviews recommend discontinuation between 34 and 36 weeks PMA, once respiratory status is stable. The infant has been apnea-free for at least five to eight days (10,14), and in the CAP trial, caffeine was permanently discontinued at a median PMA of 34.4 weeks. (15) Real-world practice, however, diverges considerably from this benchmark. In a multicenter cohort of 81,110 preterm infants discharged from 304 neonatal intensive care units in the United States between 2001 and 2016, the mean PMA at caffeine discontinuation ranged from 32 to 37 weeks across sites (16), demonstrating that variability of this magnitude exists even within high-income countries that have established protocols.

In LMIC settings, this variability reflects limitations in access to care. A landscape evaluation across five LMICs found inconsistent caffeine availability, no national policies, and only 29% of eligible infants treated. (17) Two Regional randomized controlled trials provide the closest comparison to our practice. Prakash et al., in Kerala, randomized 120 preterm neonates (26–32 weeks gestation) to caffeine discontinuation after a 7-day apnea-free period versus continuation until at least 34 weeks PMA; recurrence of apnea was similar in both arms (15% vs 13%), with caffeine stopped at a median of 33 and 34 weeks PMA, respectively. (18) Pradhap et al. likewise evaluated the optimal timing of discontinuation and found comparable overall recurrence between groups (12.6% vs 4.6%, p=0.06). (19) Published data from Bangladesh on PMA at caffeine discontinuation are not currently available, leaving a clear evidence gap for the wider South Asian region.

Against this backdrop, caffeine discontinuation in our cohort centered on the 36.1–38.0 week PMA window, accounting for 58.5% of infants in the self-weaning group and 54.5% in the dose-adjusted group. A small subgroup within the dose-adjusted group (5.5%) continued caffeine to 41–44 weeks PMA, whereas only 1 infant (1.1%) in the self-weaning group continued past 40 weeks. This timing is later than most HIC protocols target (typically 34–36 weeks PMA in clinically stable infants) and later still than both the regional Indian benchmarks and the CAP median. (10) The rightward shift is plausibly attributable to two factors. First, discontinuation was guided by attending physician discretion rather than a standardized stopping protocol, introducing clinician-level variability that may favor a more conservative threshold. Second, caregiver anxiety in an outpatient setting with limited home monitoring likely reinforced this conservatism. Notably, despite the extended exposure, no apneic episodes were reported after discontinuation in either group.

The duration of caffeine therapy involves implications beyond apnea prevention, because caffeine also influences neurodevelopment. A recent meta-analysis found that caffeine administration significantly improves long-term motor and cognitive outcomes. (20) However, because the CAP trial discontinued caffeine at a median PMA of 34.4 weeks (IQR 33.0–35.9), its long-term safety data describe outcomes after relatively early discontinuation rather than after therapy continued into or beyond this window. (15) The effects of prolonged caffeine exposure on growth and brain development therefore remain uncertain and have not been fully excluded. Taken together, our data may support 37 weeks PMA as a defensible lower bound for caffeine discontinuation in clinically stable infants in this setting; the upper bound (whether 40 weeks, 42 weeks, or beyond constitutes meaningful overtreatment) cannot be settled here. Our primary outcomes were the proportion of infants continuing caffeine beyond 36 weeks PMA and the occurrence of caregiver-reported apnea after discontinuation, and this pragmatic design cannot establish 42 weeks PMA as a definitively safe upper bound.

### Strengths

Strengths include the prospective pragmatic design, standardized data collection, structured outpatient follow-up, and multivariable adjustment for baseline gestational age and weight imbalance. The cohort reflects real-world LMIC tertiary practice.

### Limitations

Several limitations warrant explicit acknowledgment. All apneic events after discharge were caregiver-reported, with no objective home monitoring through apnea sensors or continuous pulse oximetry. Self-limiting episodes that did not prompt an emergency department visit or follow-up call would not have been captured so that post-discharge apnea may be underreported; in particular, subclinical events such as intermittent hypoxemia detectable only by continuous saturation monitoring could not be ascertained The study was also underpowered for stratified analysis within narrower corrected gestational age bands, given the rarity of recurrence events, and therefore cannot resolve whether 37, 40, or 42 weeks PMA represents the true optimum. Because the study was observational, regimen was selected at physician discretion, raising the possibility of confounding by indication. Finally, the single-center design limits external generalizability; parental discontinuation before scheduled visits (11.8% self-weaning, 9.0% dose-adjusted) introduced additional noise into the discontinuation-age distribution.

### Implications And Future Research

Overall, both self-weaning and dose-adjusted caffeine regimens appear safe in LMIC tertiary settings supported by structured neonatal clinic follow-up, and self-weaning offers a more parent-friendly and financially feasible option. A multicenter randomized controlled trial comparing discontinuation at 36 versus 40 weeks PMA, incorporating home pulse oximetry and adequately powered to detect rare apneic recurrences while minimizing risk of bias, is needed to definitively resolve the question of optimal discontinuation timing in resource-limited contexts.

## CONCLUSION

Clinically observed outpatient outcomes did not differ between infants managed with self-weaning and those managed with dose-adjusted caffeine regimens, with no reported post-discontinuation apnea in either group, very few documented non-specific caffeine-related adverse effects at home, and low apnea-related readmission rates. The findings support the feasibility of self-weaning as a pragmatic outpatient approach within structured follow-up systems, while also highlighting the need for future multicenter studies incorporating objective monitoring and strategies to reduce missing data to strengthen causal inference and generalizability.

## Data Availability

All data produced in the present study are available upon reasonable request to the authors.

